# Refined characterization of circulating tumor DNA through biological feature integration

**DOI:** 10.1101/2021.08.11.21261907

**Authors:** Havell Markus, Dineika Chandrananda, Elizabeth Moore, Florent Mouliere, James Morris, James Brenton, Christopher G. Smith, Nitzan Rosenfeld

**Author notes:** Amsterdam UMC, Vrije Universiteit Amsterdam, Department of Pathology, Cancer Centre Amsterdam, 1081 HV, Amsterdam, The Netherlands. Pennsylvania State College of Medicine, 700 HMC Cres Rd, Hershey, PA 17033. These authors contributed equally to this work (Havell Markus, Dineika Chandrananda). These authors jointly supervised this work (Christopher G. Smith, Nitzan Rosenfeld). **Author contributions:** Concept and design of the study: H.M., D.C., C.G.S. and N.R.; Methodology: H.M., D.C., F.M. and J.M.; Investigation: H.M. and D.C.; Sample Collection: E.M., J.B., and C.G.S.; Data Analysis: H.M. and D.C. Writing: H.M., D.C., F.M., C.G.S and N.R.; Funding Acquisition: C.G.S. and N.R. Supervision: D.C., C.G.S. and N.R. **Competing interest statement:** N.R. and J.B. are co-founders, present/former officers or consultants and/or shareholders of Inivata Ltd, a cancer genomics company that commercialises circulating DNA analysis. C.G.S. has consulted for Inivata Ltd. Inivata had no role in the conception, design, data collection and analysis of the study. Other co-authors have no conflict of interests.

## Abstract

Circulating tumor DNA (ctDNA) in blood plasma is present at very low concentrations compared to cell-free DNA (cfDNA) of non-tumor origin. To enhance ctDNA detection, recent studies have been focused on understanding the non-random fragmentation pattern of cfDNA. These studies have investigated fragment sizes, genomic position of fragment end points, and fragment end motifs. Although these features have been described and shown to be aberrant in cancer patients, there is a lack of understanding of how the individual and integrated analysis of these features enrich ctDNA fraction and enhance ctDNA detection.

Using whole genome sequencing and copy number analysis of plasma samples from 5 high grade serious ovarian cancer patients, we observed that 1) ctDNA is enriched not only in fragments shorter than mono-nucleosomes (∼167bp), but also in those shorter than di-nucleosomes (∼240-330bp) (28-159% enrichment). 2) fragments that start and end at the border or within the nucleosome core are enriched in ctDNA (5-46% enrichment). 3) certain DNA motifs conserved in regions 10bp up- and down-stream of fragment ends (i.e. cleavage sites) could be used to detect tumor-derived fragments (10-44% enrichment). We further show that the integrated analysis of these three features resulted in a higher enrichment of ctDNA when compared to using fragment size alone (additional 7-25% enrichment after fragment size selection).

We believe these genome wide features, which are independent of genetic mutational changes, could allow new ways to analyze and interpret cfDNA data, as significant aberrations of these features from a healthy state could improve its utility as a diagnostic biomarker.

**Significance:** In recent years circulating tumor DNA (ctDNA) has received much attention, and investment, as a biomarker that could transform the clinical care of cancer patients. Despite this, there is much that is not known about this biomarker. Recently, it has been demonstrated that the biological properties of ctDNA can be leveraged to improve ctDNA based assays. Here we build on this by carrying out an in-depth analysis of three genome wide fragmentation patterns of cell-free DNA; specifically fragment size, positioning of fragment end points with respect to nucleosome occupancy, and fragment end motifs. Whilst previous studies have described these features in an individual manner and used them as point statistics for comparison between healthy individuals and cancer patients, our study is the first to show that the individual and integrative analysis of these features can be used to enrich for ctDNA and enable enhanced ctDNA detection. The features described are independent of specific genomic alterations, with signal integrated across the breadth of the genome. This allows informative analysis that leverages a larger proportion of sequencing reads, further building the case for the use of cost-effective approaches like shallow whole genome sequencing for cancer diagnostics.

## Introduction

The utility of circulating tumor DNA (ctDNA) in plasma is well documented^1^. It is a specific biomarker that enables early detection of cancer^2^, allows treatment monitoring^3^ and provides prognostic information about tumor burden post treatment or surgery^3^. The ability to access ctDNA non-invasively is a considerable advantage over needle biopsies and allows for more frequent patient monitoring.

Despite its virtues, detecting ctDNA in blood can be likened to finding a needle in a haystack; Tumor DNA is only present in very low concentrations compared to cell-free DNA (cfDNA) of non-tumor origin, ranging from ≥5-10% in late stage to ≤0.01-0.1% in early stage cancers or early post-surgical tumor recurrence^4^. The ctDNA fractions in these early settings are often below the limit of detection of current methods.

Several studies have highlighted that ctDNA fragments tend to be shorter (around 50-150bp) than cfDNA^5–7^, and selecting these fragments could provide a median two-fold enrichment^5^. However, there is still no consensus on which fragment sizes are the most informative, and no confirmed biological explanation as to why ctDNA is shorter. Other features of cfDNA have been identified that could enrich for ctDNA including, for example, fragment end motifs^8^ and ‘preferred ends’^9^, and genomic localisation^10^. Whilst strides are being made to explore the mechanisms behind these features^11–13^, much work remains to understand how to best leverage them for improved ctDNA, and disease, detection.

cfDNA is released into the bloodstream from various cellular origins^14^ and by different biological processes such as apoptosis or necrosis^15^. This is supported by the fragment size distribution of cfDNA, which shows a modal size of 167bp corresponding to DNA wrapped around histone (∼147bp) plus linker region (∼10bp). The different ancestries can mark cfDNA with specific nonrandom fragmentation features that could be used to trace the molecules back to their origin or classify plasma samples as healthy or cancer^16^.

We hypothesized that characterization of additional distinctive fragmentation features could further improve sensitivity for detection of ctDNA as it will maximize signal to noise ratio by enriching for tumor derive fragments and filtering out wild type fragments. We believe such genome wide features, that are independent of genetic mutational information, could either be used prior to sequencing or post sequencing before variant calling, or as confidence measures, or statistical weights for anomaly detection of ctDNA amongst cfDNA.

To evaluate this hypothesis, we conducted a comprehensive assessment of molecular and fragmentation features existing in DNA fragments from plasma of high grade serious ovarian cancer (HGSOC) patients in contrast to healthy controls. We describe three biological features that are enriched in tumor-derived DNA and investigate their utility to improve ctDNA detection.

The utility of these features was assessed by first carrying out an in-silico selection of fragments that carried the feature of interest, down sampling them to mimic shallow whole genome sequencing (sWGS; 0.1X genome coverage), and measuring their respective ctDNA fraction based on copy number aberration (CNA) analysis. Plasma samples from HGSOC patients were chosen since HGSOC is mainly driven by highly abnormal copy number (CN) and structural variation (SV). Furthermore, besides alterations in the TP53 gene, very few ubiquitous point mutations are found in HGSOC. Thus, changes in ctDNA fraction in WGS data from HGSOC patients are better estimated through changes in CNA levels.

## Results

### Baseline TP53 mutant allele fraction and cancer cell fraction measurements

In order to determine what molecular features are differentially enriched in plasma cfDNA originating from healthy cells and ctDNA originating from tumor cells, paired-end whole genome sequencing was carried out on plasma samples from 5 HGSOC patients with 22.75-35.61X genomic coverage. A pool of controls was constructed by merging paired-end reads from sWGS (mean 0.4X genomic coverage) of 49 plasma samples from healthy volunteers (total 46.46X genomic coverage after merging). Thus, we had a total of 6 WGS datasets (5 patients and 1 control).

The TP53 mutant allele fraction (MAF) was measured for the plasma samples from 5 HGSOC patients using targeted amplicon sequencing (See Table S1 for amplicon details). In addition, an overall tumor fraction of the plasma samples from 5 HGSOC patients and the pool of controls was measured using ichorCNA^17^. In order to simulate sWGS and obtain a robust tumor fraction measurement, we created 10 random bootstrap sets (RBS) of 2.2 million fragments (0.1X genomic coverage) for each the of the 6 WGS datasets. For ichorCNA analysis, we built a panel of normals using the 10 RBS from the pool of controls, and measured the tumor fraction of all RBS using it as a reference.

ichorCNA estimated a tumor fraction of 0 for all 10 RBS from the pool of controls. However, in 3 out of 5 HGSOC patients, the estimated tumor fraction was lower than the TP53 MAF determined by amplicon sequencing (Figure S1). This could be due to variable copy number of the TP53 gene present across samples, which would affect the TP53 MAF i.e. the tumor fraction measured using copy number analysis is based on genome wide events, which might differ from the allele frequency of point mutations especially if they are not in copy number neutral regions. The TP53 copy number was neutral in Patient1 and the tumor fraction measured using ichorCNA (0.09) was similar to the TP53 MAF (0.11) (Figure S1). Conversely, there was a loss of the region containing TP53 in Patient2 and the tumor fraction measured using ichorCNA (0.324) was greater than the TP53 MAF (0.22) (Figure S1).

### Fragmentation feature generation

For each of the 6 WGS datasets, every properly paired read was summarized as a single fragment. Each individual fragment was annotated with 4 molecular features: fragment size, position of fragment start and end position with respect to nearest nucleosome dyad, and per-base-tri-nucleotide-bias score (See Methods for more details).

The capacity of each feature to select for cfDNA or ctDNA was measured by selecting fragments with a given feature cutoff, creating 10 RBS of 2.2 million fragments for all 6 WGS datasets, constructing a panel of normal using the 10 RBS from the pool of controls, and using it as a baseline for measuring their tumor fraction using ichorCNA. We created a different panel of normals for each feature and applied a cutoff to reduce noise and correct for systematic biases that may arise from library construction, sequencing platform, cfDNA specific artifacts, and feature selection. This process, which we refer to as ctDNA in-silico bootstrap enrichment process (CISBEP), was carried out for all features with various cutoffs. To determine which cutoff resulted in the highest enrichment of tumor fragments, we calculated the relative tumor fraction, which was the ratio of the median tumor fraction from the 10 RBS of fragments within cutoff to the median tumor fraction of fragments with no cutoff or no selection. An overview of the study and CISBEP is illustrated in Figure 1.

**Figure 1.**
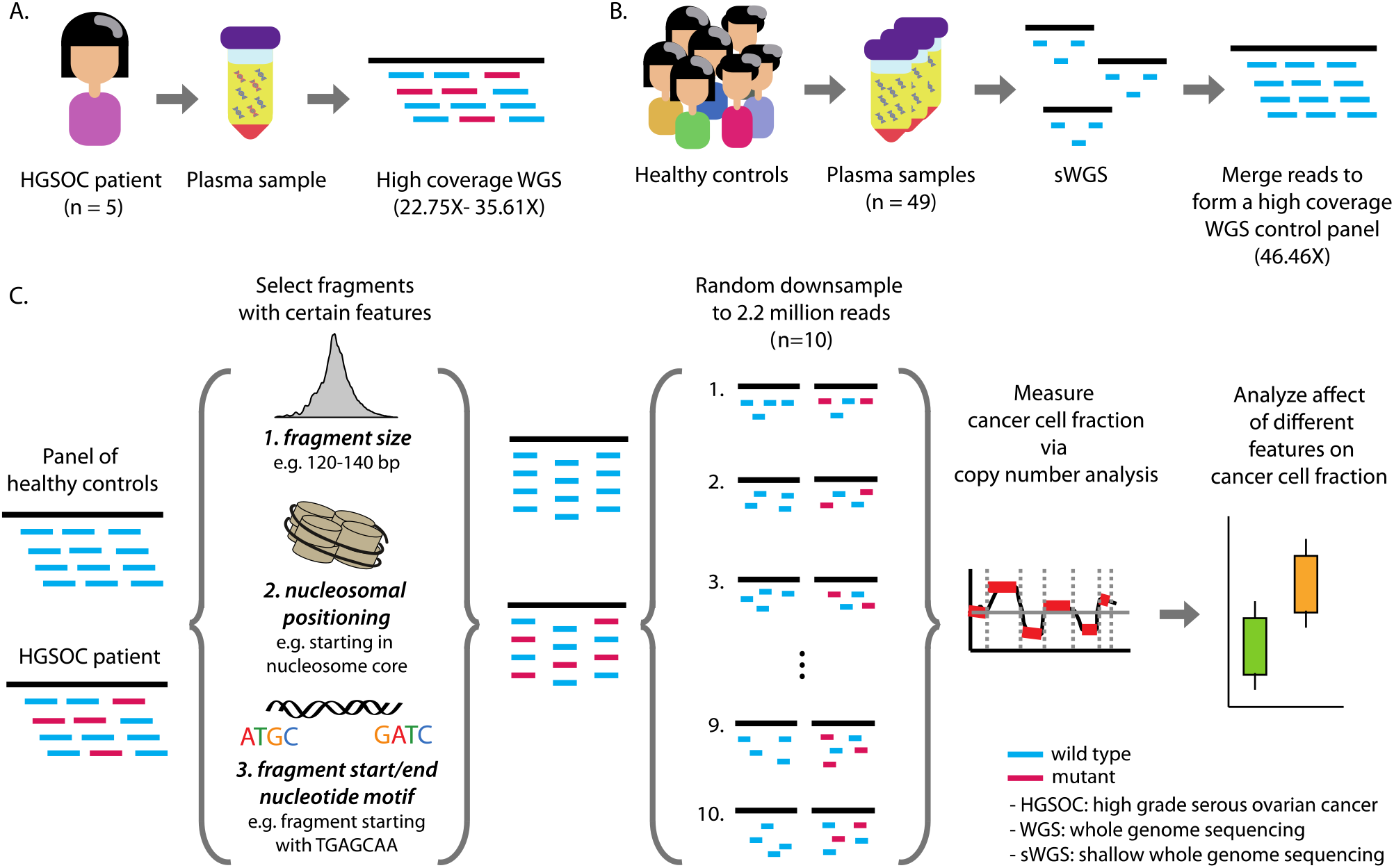
(A) In this study we performed paired-end high coverage whole genome sequencing on plasma samples from 5 high grade serous ovarian cancer patients and (B) paired-end shallow whole genome sequencing (sWGS) on 49 healthy plasma controls. We created a control panel by merging reads across all healthy samples (C) Three features were calculated for each plasma DNA fragment: fragment size, relative position of fragment start and end sites with respect to the closest nucleosome dyad, and a nucleotide frequency score based on the 10 bp region spanning either side of fragment start and end. For both the control panel and the patient high-coverage datasets, fragments were selected based on these features and repeated random down-sampling was carried out in order to create 10 technical replicates (0.1x coverage, 2.2 million reads each). These shallow whole-genome datasets underwent copy number analysis and the cancer-cell fraction (CCF) was calculated using ichorCNA. This allowed us to determine features that are enriched in circulating tumor DNA versus circulating cell-free DNA.

### Fragment size

We created fragment size bins based on the ∼10bp periodic peaks in the range 0-200 bp and ranges where the position of the di-nucleosome peak showed a disparity between the panel of normals and cancer patients (Figure 2A). This resulted in a total of 16 fragment size bins (0-52bp, 53-64bp, 64-73bp, 74-83bp, 84-94bp, 95-104bp, 105-114bp, 115-125bp, 126-135bp, 136-143bp, 144-153bp, 154-160bp, 161-171bp, 172-239bp, 240-324bp, and 325-400bp). Fragments within the ranges of these bins were selected, and for each fragment size bin 10 RBS of 2.2 million fragments were created for all 6 WGS datasets. The enrichment of ctDNA within each fragment size bin was then measured through the CISBEP.

**Figure 2.**
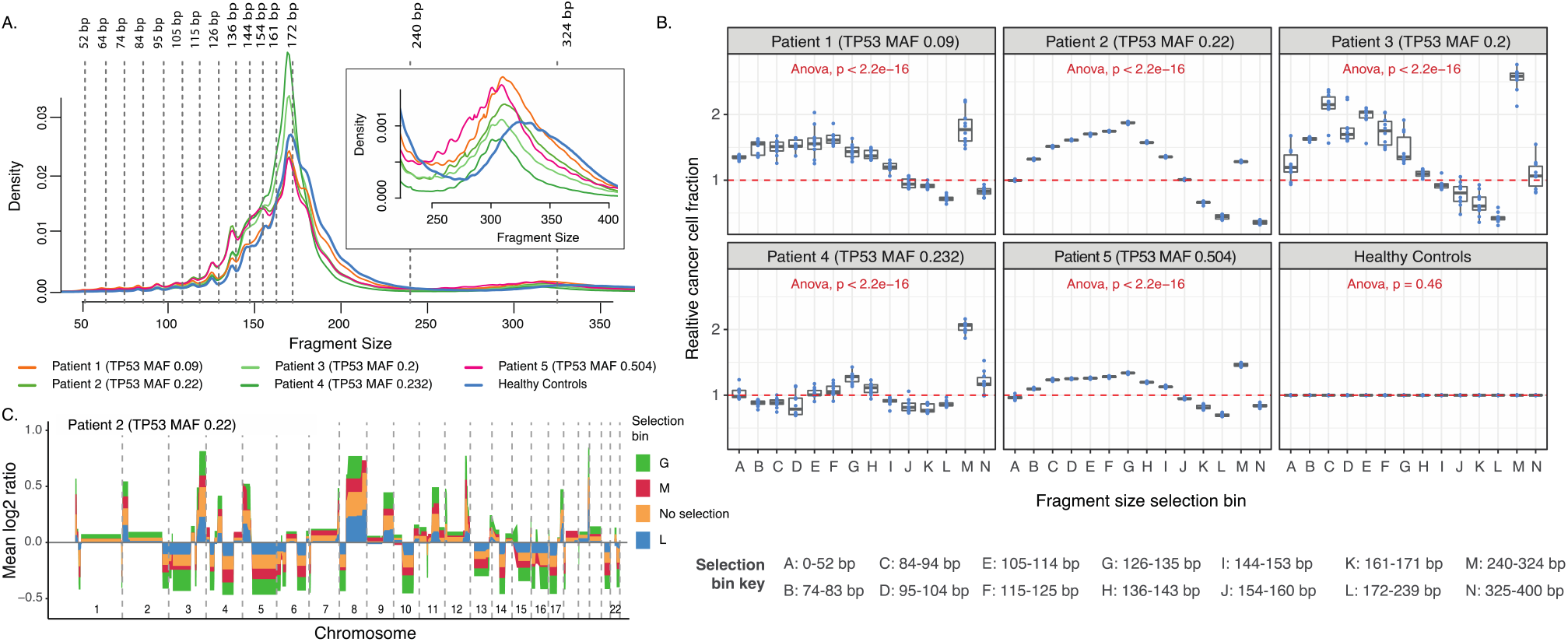
Fragment size analysis. (A) Fragment size distribution of 5 HGSOC patients and panel of healthy controls. The vertical dashed lines are placed on the fragment sizes between 52 and 172 bp where 10 bp periodicity is observed. The vertical lines at 240 and 324bp show the range at which a shift in the di-nucleosomal peak occurs between HGSOC patients and healthy controls. The inset plot enlarges the density profile in the di-nucleosomal fragment length range. (B) Copy number aberration analysis of all technical replicates comprising of different fragment length ranges as indicated by figure legend. The cancer cell fraction (CCF) was measured for each dataset with and without fragment selection. The relative CCF on the y-axis is the ratio of these two estimates. Above each plot, the TP53 MAF, as determined by TAm-Seq is shown for each patient (C) The average relative copy number values from the 10 replicates of patient 2. The different colors indicate different size selection ranges as well as no size selection. Corresponding data from the other patients are shown in supplementary figure 3.

Figure 2B shows the relative tumor fraction of all 10 RBS from the 6 WGS datasets for all the fragment size bins. As bins 53-64bp and 64-73bp contained fewer than 2.2 million fragments in the pool of controls, they were not considered for further analysis. The relative tumor fraction remained at 0 in all bins for the panel of controls, indicating that fragment sizes of variable lengths did not change the genome wide copy number in healthy plasma samples. Conversely, the relative tumor fraction was significantly different across all bins in the 5 HGSOC patients (ANNOVA p-value < 2.2e-16). Of these, the fragments within 126-135bp and/or 240-324bp bins consistently showed the highest enrichment of tumor fraction across all 5 HGSOC patients as their median relative tumor fraction ranged from 1.28-1.87 and 1.28-2.59 respectively. This meant ctDNA enrichment of 28-87% and 28-159% in fragments of sizes 126-135bp and 240-324bp respectively.

The relationship between the fragment size bins and ctDNA enrichment was noticeable in log2 ratios of copy number analysis, as the regions with copy number aberrations deviated further from or closer to copy number neutrality. Figure 2C illustrates the increased deviance of copy number aberrations in each fragment size bin, especially in bins 126-135bp and 240-324bp for Patient 2. This observation was similar across the remaining 4 HGSOC patients (Supplementary Figure 3).

We further observed that fragments within the 154-160bp, 161-171bp, and 172-239bp bins consistently resulted in relative tumor fractions lower than 1, in-turn suggesting that fragments with these sizes are enriched for wild type or healthy cfDNA (i.e. non-mutant tumor derived DNA or DNA originating from healthy cells).

### Fragment start and end position relative to nucleosome center

We next hypothesized that, if cfDNA fragments are protected from enzymatic degradation through association with nucleosomes, then perhaps the shorter fragments that are enriched in ctDNA are frequently being cleaved closer to the nucleosome boundary or core, as previously shown in fetal cfDNA^18^. To evaluate this, we downloaded the CH01 genome-wide nucleosome track, as used by Snyder et. al.^16^ which contains 12.9 million nucleosome calls inferred from the in-vivo positioning of plasma cfDNA. We annotated the relative positioning of start and end sites of all fragments in the 6 WGS datasets with respect to the nearest nucleosome center from the CH01 track (See methods for details). Consistent with our hypothesis, fragments from the 5 HGSOC patients show a higher density of start and end positions within 75bp downstream and upstream of the nucleosome dyad when compared the fragments from the pool of controls (Figure 3A).

**Figure 3.**
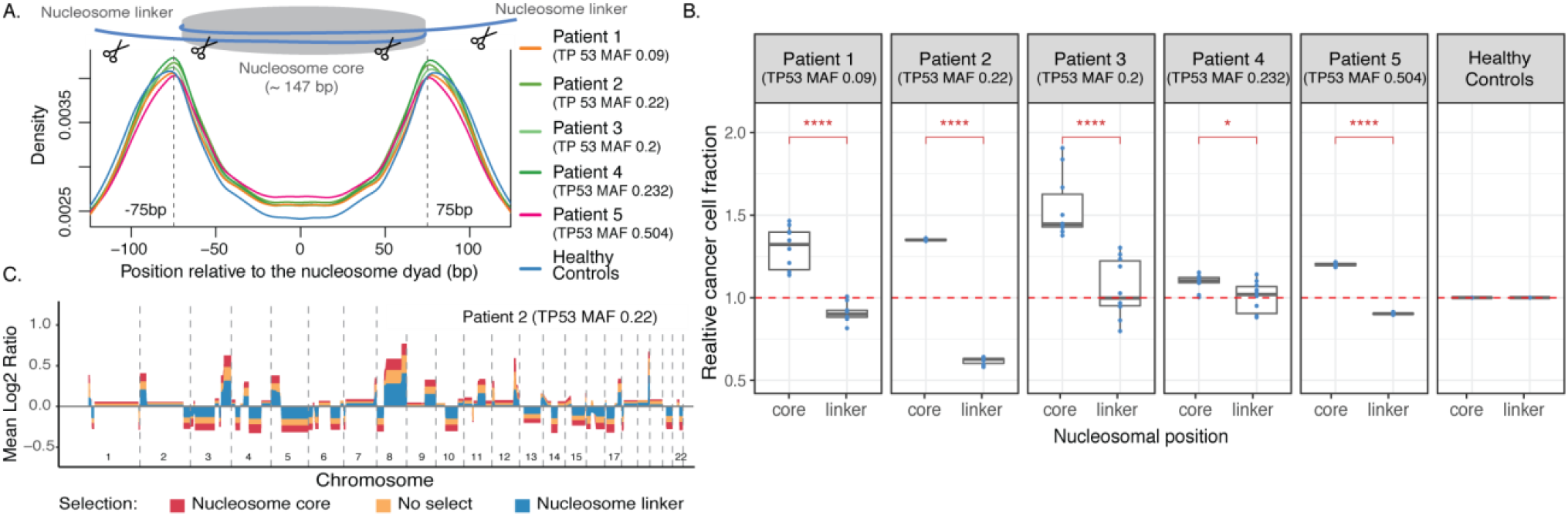
Location of DNA fragments relative to nucleosomal dyads. (A) The density of fragment start and end sites with respect to nucleosome dyads for patients and controls. The vertical lines are drawn at 75 bp downstream and upstream of the nucleosome center to signify the region of nucleosome core and linker. (B) Copy number aberration quantification of all technical replicates containing fragments starting and ending within nucleosome core or linker regions. (C) The average relative copy number values from the 10 replicates of patient 2. The different colors indicate results from using fragments that start and end within nucleosome core region, and fragments that start and end within nucleosome linker region as well as all fragments with no selection.

To investigate whether fragment positioning could be used to enrich for ctDNA, we selected fragments that start and end within (nucleosome core region) and outside (nucleosome linker region) of 75bp downstream and upstream from the nucleosome dyad, and evaluated ctDNA enrichment through the CISBEP for the 6 WGS datasets. We observed fragments that start and end in the nucleosome core tend to be shorter, and this is observed even in the di-nucleosome peak, as it is shifted towards shorter fragment size (Figure S4).

In all 5 HGSOC patients, the relative tumor fraction of the fragments that started and ended within the core of the nucleosome was significantly higher than the fragments that started and ended within the linker region (t-test, p-value < 0.0001-0.05) (Figure 3B). In addition, fragments in the linker region consistently resulted in relative tumor fraction lower than 1, which suggests these fragments are enriched for wild type or healthy cfDNA. The relative tumor fraction of fragments that started and ended in the nucleosome core ranged from 1.1-1.44 (i.e. a 10-44% enrichment of ctDNA). Figure 3C illustrates the increase in the level of copy number aberration levels affected by fragments that start and end within nucleosome core as compared to the linker region for Patient 2. This observation was similar across the remaining 4 HGSOC patients (Figure S5). Taken together, these observations suggest fragments starting and ending within nucleosome core are enriched in ctDNA, while fragments starting and ending in linker regions are enriched in wild-type cfDNA.

### Fragment start and end flanking 10bp tri-nucleotide frequency motif

Previous studies have shown that nucleosome positioning is sequence dependent, which varies between core and linker regions^19^. Chandrananda et al. showed DNA motifs in regions 10bp downstream and upstream of cfDNA cleavage sites are consistent and conserved in healthy controls^20^. Thus, we hypothesized there might be different nucleotide motifs present in fragment start and end sites of cfDNA and ctDNA as they have altered nucleosome positioning. To investigate whether such differences exist, we investigated the per base mono-, di-, and tri-nucleotide frequencies in regions 10bp downstream and upstream of fragment start and end sites from the pool of controls and HGSOC Patient 5 (the patient with highest TP53 MAF of 0.504) (Figure S6-11). Although the per-base-nucleotide frequencies were similar between the two datasets, we found apparent differences at specific positions.

In order to leverage the fragmentation motif differences for ctDNA enrichment, we developed a computational approach to assign individual fragments a per-base-nucleotide-bias score (PBNB-score) to indicate whether a fragment is likely derived from tumor or healthy cells. The PBNB-score is based on a top-down approach, where we learned from sample level differences and applied them to discriminate individual fragments. To learn sample level differences, we generated 10 RBS of 3 million fragments from both the pool of controls and HGSOC Patient 5. For each of the RBS we calculated two position weight matrix (PWM) that described the per-base-mono-nucleotide-frequencies in regions -10bp and +10bp of fragment start and end sites respectively. We vectorized the PWM of fragment start sites from all the RBS and combined them into a single matrix. Similar steps were carried out for PWM of fragment end sites from all RBS. We then carried out principal component analysis (PCA) on the two matrices, which summarized the per-base-mono-nucleotide-frequencies at fragment start and end sites for all 10 RBS from both the pool of controls and HGSOC Patient 5. We carried out similar steps for per-base di- and tri-nucleotide frequencies. Figure S12 illustrates the conceptual framework of these steps.

We found that principal component 1 (PC1) of fragment start and end per base mono-, di-, and tri-nucleotide frequency matrices separated all of the 10 RBS of the pool of controls from HGSOC Patient 5 (Figure S13A). We observed the positive components of the PC1 eigenvector identified the base specific nucleotide motifs enriched in the pool of controls, while the negative components identified the base specific nucleotide motifs enriched in HGSOC Patient 5. We used the PC1 eigenvector of fragment start and end per base mono-, di-, and tri-nucleotide frequency matrix as weights to compute per-base mono-, di-, and tri-nucleotide bias score (PBMNB-score, PBDNB-score, and PBTNB-score) for each individual fragment respectively (Figure S13B and Methods for more details). The higher the score, the more likely a fragment was derived from healthy cells, while the lower the score, the more likely it was derived from the tumor cells (Figure S13C).

To determine which of the three nucleotide motif scores provided the most discrimination between cfDNA and ctDNA fragments, the Kolmogorov-Smirnov test was performed on the three score distributions of all fragments derived from the pool of controls and HGSOC Patient 5. The distribution of all three nucleotide motif scores were significantly different between the two sample types (p-value < 2.2e-16, data not shown). However, the distribution of the tri-nucleotide motif score had the largest test statistic or difference between the score distribution of the two sample types. The test statistic comparing the PBMNB-score, PBDNB-score, and PBTNB-score from the two sample types were 0.10673, 0.12738, and 0.14129 respectively. The PBTNB-score for all fragments from 5 HGSOC patients were lower than the pool of controls (Figure S13C).

We next used CISBEP to evaluate whether the PBTNB-score could be used to enrich for ctDNA by selecting fragments with a PBTNB-score ≤ and ≥ to -0.3, -0.15, 0, 0.15, and 0.3 from the 6 WGS datasets. In all 5 HGSOC patients, fragments with an increasingly negative PBTNB-score showed an increase in relative tumor fraction, while fragments with an increasingly positive PBTNB-score showed a decrease in relative tumor fraction (Figure 4A). This trend was significant in 4 out of 5 patients (ANNOVA p-value << 0.01). For fragments with PBTNB-score ≤ -0.3, the median relative tumor fraction ranged from 1.05-1.46 (5-46% enrichment of ctDNA). In contrast, for fragments with PBTNB-score ≥ 0.3, the median relative tumor fraction ranged from 0.71-0.91 (0.09-0.29% enrichment of cfDNA). Figure 4B illustrates the increase in the level of copy number aberration levels affected by fragments with PBTNB-score ≤ -0.3 and ≥ 0.3 for HGSOC Patient 2. This trend was similar across the remaining 4 HGSOC patients (Supplementary Figure 14). We also observed that fragments with PBTNB-score ≤ -0.3 tend to be shorter and fragments with PBTNB-score ≥ 0.3 tend to be longer (Figure 4C and Figure S15). The trend was stronger in HGSOC patients compared to the pool of controls. This could suggest an underlying fragmentation mechanism that is present even in healthy individuals but is enhanced in HGSOC patients. Taken together, these observations suggest that the nucleotide motif flanking 10bp position of fragment start and end sites are unique to wild-type cfDNA and ctDNA fragments, and could be used to enrich for ctDNA or filter out cfDNA fragments.

**Figure 4.**
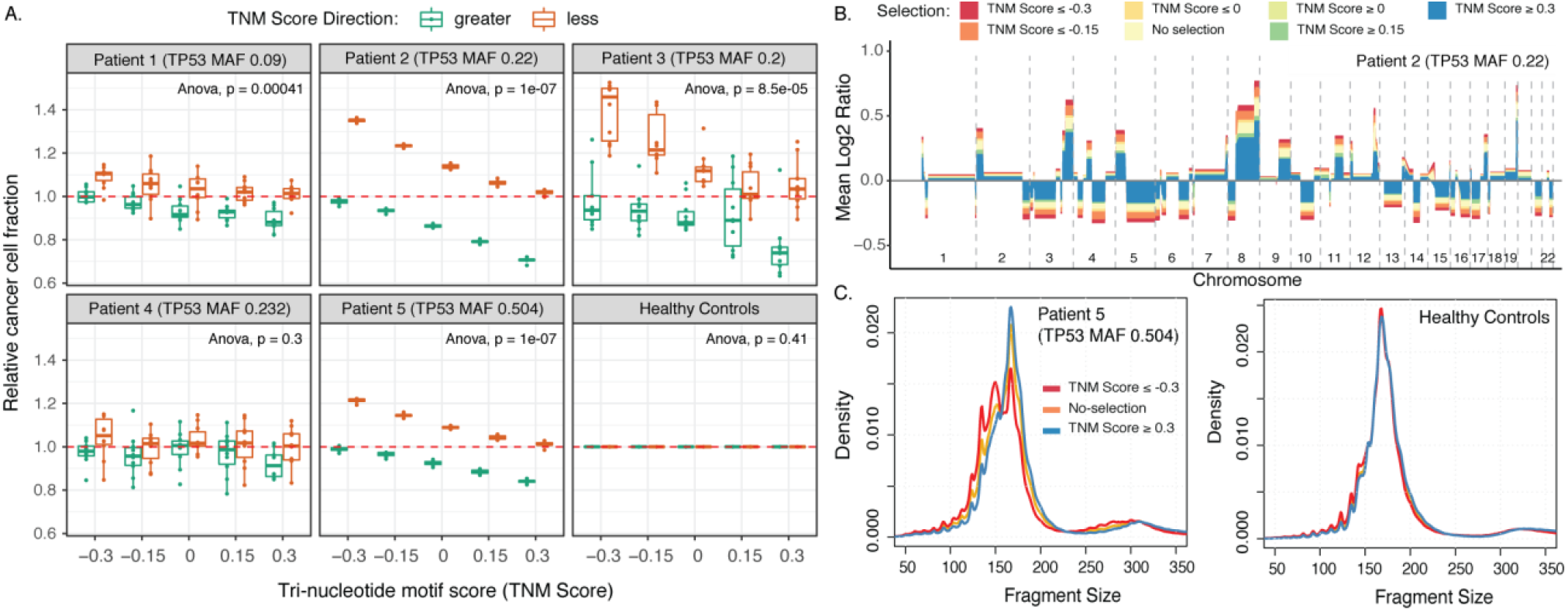
Tri-nucleotide motif score. (A) Copy number aberration analysis of all technical replicates containing fragments with tri-nucleotide motif scores less than or greater than - 0.3, -0.15, 0, 0.15, and 0.3. The CCF of all the random samples is shown relative to median CCF from unselected data. (B) The average relative copy number values from the 10 replicates of patient 2. The different colors indicate results from using fragments with no selection, fragments with tri-nucleotide motif score less than or equal to -0.3, -0.15, 0, and greater than or equal to 0, 0.15, 0.3. (C) The fragment size distribution in the control pool and HGSOC patient 5 of fragments with no selection, fragments with tri-nucleotide motif score less than or equal to -0.3 and greater than or equal to 0.3.

### Integrating fragment size and tri-nucleotide motif for ctDNA enrichment

In-silico selections of short fragments and fragments with negative PBTNB-score independently show an enrichment of ctDNA (Figure 2 and 4). To investigate whether combining these features would result in further enrichment of ctDNA, we selected three types of fragments from all 6 WGS datasets: fragments with sizes 74-144bp, fragments with sizes 74-144bp PBTNB-score ≥ 0.2, and fragments with sizes 74-144bp and PBTNB-score ≤ - 0.2. We chose a wide range of shorter fragments and cutoff of ± 0.2 PBTNB score to increase the diversity of fragments and capture genome wide variations when choosing random samples for CISBEP, and allow at least 2.2 million fragments after various layers of feature selection. We also selected fragments based on start and end position relative to nucleosome center, and applied additional PBTNB-score selection. However, due to limited number of annotated nucleosomes over the entire genome, it led to limited fragments available for CNA analysis after PBTNB-score selection. Thus, we did not consider nucleosome positioning for further feature integration analysis, and relied on fragment size as a reliable representation of fragments originating from different nucleosome positioning.

The relative tumor fraction was calculated using the CISBEP. In all 5 HGSOC patients, fragments with sizes 74-144bp and PBTNB-score ≤ -0.2 showed a significant increase in the relative CCF when compared to fragments with sizes 74-144bp (t-test, p-value < 0.01, Figure 5A). We measured the additional ctDNA enrichment by taking the fraction of median relative CCF of fragments with sizes 74-144bp and PBTNB-score ≤ -0.2 to median relative CCF of fragments with sizes 74-144bp alone. The fraction of ctDNA enrichment ranged from 1.07-1.25. This meant additional 7-25% ctDNA enrichment from applying PBTNB selection after size selection. Meanwhile, fragments with sizes 74-144bp and PBTNB-score ≥ 0.2 showed a significant decrease in the relative CCF when compared to fragments with sizes 74-144bp in 3 of 5 HGSOC patients (t-test, p-value < 0.05, Figure 5A). This suggests that combining the PBTNB-score with fragment size does provide further enrichment of ctDNA or filtering out wild-type cfDNA.

**Figure 5.**
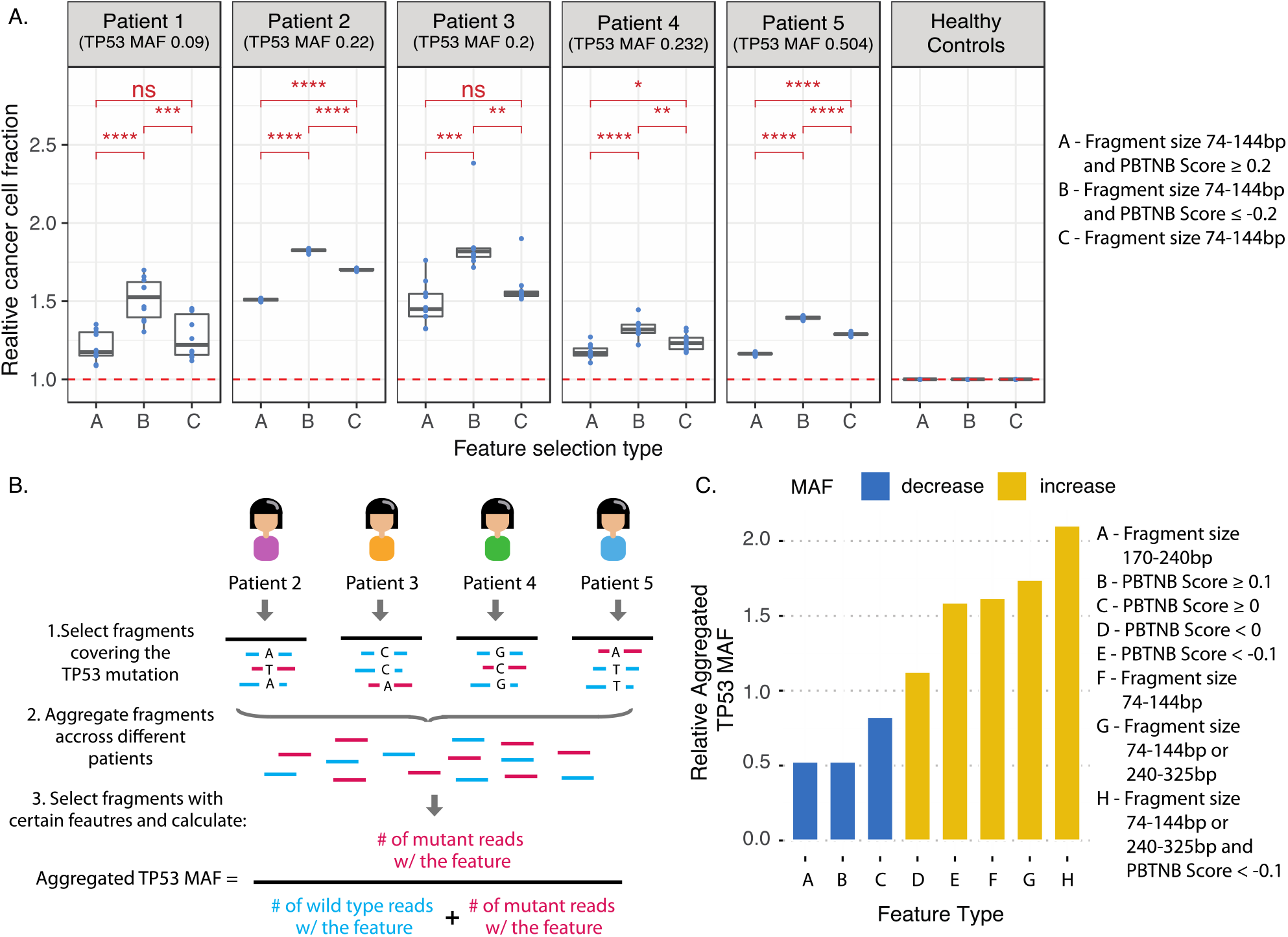
Analysis of fragments selected for multiple features. (A) Copy number aberration analysis of all technical replicates containing fragments with different lengths and motif score combinations. (B) Aggregated TP53 mutant allele fraction measurements after selecting fragments with various features. The fragments from Patient 2, Patient 3, Patient 4, and Patient 5 were aggregated as these samples had a single point mutation while Patient 1 had an insertion. (C) The aggregated TP53 mutant allele fraction is shown relative to the aggregated MAF with no-selection for different fragment feature selections.

### Effect of fragmentation feature selection on TP53 MAF

In order to further validate the utility of these features to enrich for ctDNA, we aggregated all fragments aligning to the individual TP53 point mutation locus of HGSOC Patient 2-5 (Patient 1 had an insertion). All fragments were annotated with fragment size and PBTNB-score. We did not investigate positioning of the fragments relative to the nucleosome center since some TP53 point mutations did not overlap with nucleosomes inferred from CH01. We then calculated an aggregated TP53 MAF by selecting fragments based on certain features. Figure 5B illustrates these steps. We further calculated a relative aggregated TP53 MAF by taking the ratio of the aggregated TP53 MAF with feature selection to no feature selection.

We observed that selecting fragments with sizes 170-240bp, PBTNB-score ≥ 0.1, and PBTNB-score ≥ 0 resulted in a relative aggregated TP53 MAF of 0.53, 0.53, and 0.82 (Figure 5C). This was expected as these features were related to the enrichment of wild-type cfDNA. We also observed that selecting fragments with PBTNB-score < 0, PBTNB-score < -0.1, sizes 74-144bp, and sizes 74-144bp or 240-325bp led to a continual enrichment of ctDNA fragments with relative aggregated TP53 MAF of 1.13, 1.59, 1.62, and 1.74 respectively. Combining all features, fragments with PBTNB-score < -0.1 and size 74-144bp or 240-325bp, led to the highest enrichment of ctDNA with the relative aggregated TP53 MAF of 2.10. We picked these feature cutoffs as it allowed the aggregated depth of at least 25 (range 26-71), except for aggregated depth of 12 after combination of all features (PBTNB-score < -0.1 and size 74-144bp or 240-325bp). These results provide additional validation of these features to enrich for ctDNA, as they not only led to increased tumor fraction in copy number analysis but also increased MAF in variant analysis.

## Discussion

Features described in this study are novel and could aid in the underlying biology behind origin of ctDNA in HGSOC patients. We show individual and integrative analysis of fragment size, positioning of fragment ends with respect to nucleosome center, and fragment end motif can be leveraged in fragment-specific fashion to allow enrichment of ctDNA in HGSOC patients. Although this proof-of-concept study was performed on a small sample size, we believe these features hold a great potential to guide future cfDNA fragmentomics studies and need to be validated in a large-scale cohort with multiple cancer types.

Previous studies focused on circulating DNA fragment size have used selection of short fragments (90-150 bp) for enrichment of ctDNA^3^ or differences in fragmentation size distribution across a tiled genome for cancer detection^21^. In this study, we conduct a resolute evaluation of the relative enrichment of ctDNA at each of the 10bp periodic peaks and show fragments that are shorter than both the mono- and di-nucleosome peak are similarly enriched in ctDNA. Other studies have also defined specific nucleotide motifs found at circulating DNA fragment ends that are enriched in patients with hepatocellular carcinoma and could be used as another diagnostic marker^8^. In this study, rather than looking at enrichment of certain motif at fragment ends, we use the differences in the per-base background distribution of tri-nucleotide frequencies around fragment ends of healthy controls and HGSOC patients to define a scoring system for individual fragments. We show PBTNB-score helps distinguishing fragments originating from tumor or healthy cells. Lastly, no previous study has shown a direct relationship between CCF and positioning of fragment ends to nucleosome center in cancer patients. We show that fragments starting and ending within the nucleosome core boundary are enriched in ctDNA (increased CCF), and contrastingly, fragments starting and ending in the linker region are enriched in cfDNA (decreased CCF). It is important to note that selection of these features on healthy controls had no effect of CCF, suggesting these features are biologically imprinted on fragments from their origins, rather than driven by technical artifacts.

Thus, unlike other studies, rather than defining these features as a point summary per sample for cancer detection, we show how these features provide fragment level information that could help differentiate fragments originating from tumor cells or wild type healthy cells. We show pre-selecting fragments for ctDNA analysis could maximize signal to noise ratio by either selecting fragments that are more likely to originate from tumor cells, or filtering out fragments that are more likely to originate from healthy cells. These features may also be used in confidence measures or statistical weights for genomic aberrations in order to classify them as somatic or not. They could help design fragmentation aware assays that would be more accurate and sensitive (e.g. PCR amplicons designed based on nucleosome positioning and nucleotide frequencies). This allows us to overcome the limitation of detecting a needle in the haystack, by either removing some of the hay or increasing the number needles.

Apart from limited sample size and application of these features to different cancer types, there are other limitations of this study that must be acknowledged. We used a nucleosome track that was constructed using plasma cfDNA positioning, which mainly represents hematopoietic cells^4^. This could bias the selection of fragments originating from healthy cells. Thus, selecting fragments based on a nucleosome track that is constructed using the tumor tissue or from the patients’ own plasma sample would be ideal to reduce some of the technical biases. However, we still observed enrichment of CCF, suggesting conservation of nucleosome positioning of hematopoietic and cancer cells. The weights used for calculating mono-, di-, and tri-nucleotide scores were obtained based on motif frequencies at fragment ends. Various pre-analytical and sequencing variability could influence these motifs. Future works should address the effect of these confounders to fragment end motif. The effect of pre-analytical and sequencing variability was controlled in this study as these samples were processed in a similar manner and sequenced on the same machines. Lastly, CCF measurements from ichorCNA were used as a metric to quantify the relationship between fragments with certain features and their relative enrichment of ctDNA. However, the CCF measurements could be inaccurate as the algorithm may choose a suboptimal solution. This was noted in some cases as a large portion of log2 ratios were labeled as gains or losses which suggested whole genome amplification event or apparent CNAs were called as neutral. This led to increased variability of the measured CCF. The developers of ichoCNA suggest manually checking other solutions based on ranked log likelihood to see if any provide a better explanation of the data. However, manual screening is not feasible when quantifying hundreds of samples.

In summary, this study describes three biological features that could be used to select fragments originating from tumor cells or filter out fragments originating from healthy cells. Although there are some limitations to this study, the enrichment of ctDNA captured by these features provides biological insights about previously described and poorly understood relative shortening of ctDNA. In addition, it provides novel information about sequence motifs present in the cleavage sites of cfDNA. Lastly, it shows these features could be used to conduct selective sequencing or perform in-silico selection of cfDNA fragments to be analyzed. Many labs are now carrying out sWGS or WGS routinely on plasma samples. In addition to variant and copy number analysis, we believe bioinformatic exploration of these features could further aid and help exploit cfDNA signatures that could lead to a comprehensive and robust analysis. We believe these genome wide features that are independent of genetic mutational information could be more sensitive in identifying disease status, thus leading to creation of complex diagnostics tools that could integrate additional information.

## Materials and Methods

### Sample collection and processing

Plasma samples from 49 healthy volunteers and 5 high grade serous ovarian carcinoma (HGSOC) patients were collected. The 49 healthy volunteers used in this study where part of a previous study^5^. The plasma samples from HGSOC patients were collected at Addenbrooke’s Hospital, Cambridge, UK, approved by the local research ethics committee (REC reference number: 07/Q0106/63; and National Research Ethics Service Committee East of England–Cambridge Central 03/018). Written informed consent was obtained from all patients and blood samples were collected before initiation of treatment with surgery. DNA was extracted from 4 ml of plasma using the QIAamp Circulating Nucleic Acid Kit (QIAGEN;) or QIAsymphony (QIAGEN;) according to the manufacturer’s instructions. Baseline tumor tissue biopsies were available from all 5 HGSOC patients. All 5 HGSOC patients had TP53 mutations which were determined through tumor and germline sequencing (Table S1). The TP53 mutant allele frequency in plasma samples was subsequently determined using targeted amplicon sequencing. The libraries were prepared using a previously described method, Tagged-Amplicon Deep Sequencing^22^. Primers were designed to cover the whole coding TP53 gene, which consisted of 21 amplicons with mean size of 100bp. Libraries were sequenced using MiSeq or HiSeq 4000 (Illumina).

### Analysis of WGS data

Indexed sequencing libraries were prepared using the ThruPLEX Plasma-seq kit (Rubicon Genomics). For healthy volunteers, libraries were pooled in equimolar amounts and sequenced to <0.4x depth of coverage on a HiSeq 4000 (Illumina), generating 150-bp paired-end reads. For 5 HGSOC patients, libraries were sequenced to 22.75-35.61x depth of coverage on a HiSeq 4000 (Illumina), generating 150-bp paired-end reads. The sequenced reads were aligned to the human genome (hg19) using bwa mem v0.16a. The aligned bam files were sorted and indexed using samtools v1.7. PCR and optical duplicates were marked and removed using Picard tools v2.17.6 MarkDuplicates. Reads that were unmapped, supplementary alignments, or not a primary alignment were excluded from downstream analysis. Local realignment of reads around indels was performed using GATK v0.2.2 IndelRealigner. This process realigns the misaligned reads near known indels to minimize number of mismatched bases across all reads. Lastly, read base quality scores were recalibrated using GATK BaseRecalibrator and ApplyRecalibration to correct for systematic errors in the base quality scores produced by the sequencer. Reads with final mapping quality of less than 20 were excluded from downstream analysis. Reads from all healthy volunteers were merged into a single bam file using Picard tools v2.17.6 MergeBamAlignment to construct a pool of controls (46.46x depth of coverage). Paired end reads for all 6 WGS datasets (pool of controls and 5 HGSOC patients) were summarized as fragments with their 3’ and 5’ position into a bed file using samtools v1.7 and a custom script. The remainder of the analysis was carried out using only autosomes and fragments with size ranging from 0-800bp.

### Fragment size and nucleosome positioning analysis

For each fragment in the bed file we calculated their fragment size by subtracting the 5’ and 3’ position. We also determined the relative distance of fragment start and end sites to its nearest nucleosome center using the CH01 nucleosome track (12.9 million nucleosome calls) from Snyder et al^16^. We intersected the bed file of all fragments with CH01 nucleosome bed file in R using the GenomicRanges package. For each overlap hit, the distance of fragment start and end position from the center of the nucleosome was calculated. In cases where there was more than one overlap, the minimum relative start and end distance from the nucleosome was considered. This captures the start and end distance with respect to their closest nucleosome.

### Per-base nucleotide bias score

To evaluate the utility of the nucleotide motif flanking 10bp downstream and upstream of fragment start and end sites, we developed a method to assign each fragment a score that indicates whether a fragment is likely to originate from tumor or normal cells.

To do so, we created 10 random bootstrap sets (RBS) of 3 million fragments from the pool of controls and HGSOC Patient 5 who had the highest TP53 MAF at 0.504. For each random sample, two per base position weight matrices (PWM) were constructed from the genomic sequences flanking 10bp downstream and upstream of both fragment start and end sites. For nucleotide sequences of length 20 the PWM for mono-nucleotide frequency is a 4 × 20 matrix. Where an entry *W*_*i,n*_, *i* ∈ {*A, T, C, G*} and *n* ∈ {−10, −9, −, −1,0,1, …, 9}, is the mean frequency of *i*^*th*^ nucleotide present at position *n*. Similarly, two di- and tri-nucleotide PWM were also constructed for each RBS for both fragment start and end sites. Di- and tri-nucleotide PWM is a 16 × 20 matrix and 64 × 20 matrix respectively describing the relative frequencies of each k-mer of length 2 and 3 at each position. Thus, for each RBS, a total of 6 PWM were constructed (mono-, di-, and tri-nucleotide frequencies of sequences flanking 10bp downstream and upstream of both fragment start and end sites).

Next, we carried out principle component analysis (PCA) on each of the 6 PWM individually. We found in all 6 PCA, the principal component 1 separated all the 10 RBS from pool of controls and HGSOC Patient 5 in unique clusters. The factor loadings of PC1 indicate the correlation between the two sample types and the nucleotides motifs in the flanking 10bp downstream and upstream of both fragment start and end sites. Therefore, loadings from PC1 that point in the direction of the pool of controls correspond to the position and nucleotide motif that are more associated with the wild type fragments, while those ones pointing to HGSOC Patient 5 are associated with tumor derived fragments. In our specific case, negative and positive elements of the loadings vector pointed to HGSOC Patient 5 and pool of controls respectively. We next obtained PC1 loadings of all 6 matrices, and used them as weights to assign per-base mono-, di-, and tri-nucleotide bias scores to individual fragments. The PC1 loading vector of mono-, di-, and tri-nucleotide PWM of fragment start and end sites will be referred to as 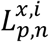, where *p* ∈ {*start, end*},

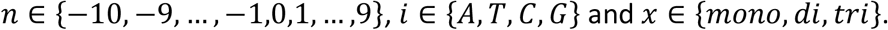

For a given fragment, the genomic sequences flanking 10bp downstream and upstream of both fragment start and end sites were obtained in R using the Biostrings package. For calculating the per-base mono-nucleotide bias score (PBMNB-score) for a fragment, the start and end sequences were summarized into two binary 80 × 1 vectors 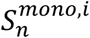 and 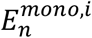 respectively, where n ∈ {™10, ™9, …, ™1,0,1, …, 9}, *i* ∈ {*A, T, C, G*}, and if nucleotide *i* exists in position *n* of start (or end) sequence the 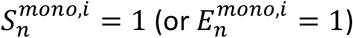 otherwise 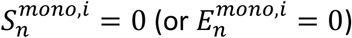. The PBMNB-score was calculated as shown in Equation 1.

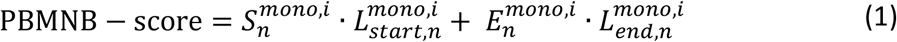

Similarly, for a given fragment, two binary 320 × 1 vectors 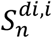 and 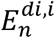 were created to describe whether a di-nucleotide motif *i* exists on position *n* of start or end sequences respectively. The per-base di-nucleotide bias score (PBDNB-score) was calculated as shown in Equation 2.

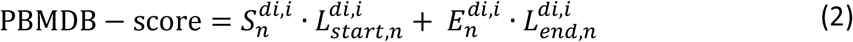

Lastly, for a given fragment, two binary 1280 × 1 vectors 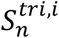 and 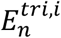 were created to describe whether a tri-nucleotide motif *i* exists on position *n* of start or end sequences respectively. The per-base tri-nucleotide bias score (PBTNB-score) was calculated as shown in Equation 3.

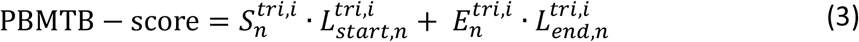

A negative motif score was hypothesized to be more associated with tumor fragments, while a positive score with normal wild-type fragments. It is important to note that depending on samples the rotations of PCA the association of negative and positive score to normal and tumor fragments might change.

### ctDNA in-silico bootstrap enrichment process (CISBEP)

After calculating the 4 molecular features for all fragments belonging to the 6 WGS datasets, we evaluated the relation of each feature to enrich for cfDNA or ctDNA. In order to do so, we selected fragments with a given feature cutoff (e.g. fragments with size ranging from 74-83bp). For all 6 WGS datasets, the selected fragments were summarized in a bam using Bedtools v2.27.1. A bam file was also created with fragments with no feature cutoff for all 6 WGS datasets (e.g. fragments with no size cutoff). These bam files will be referred to as with cutoff and no cutoff.

Once the cutoff and no cutoff bam files were created for all 6 WGS datasets for a given feature with a certain cutoff, we created 10 RBS of 2.2 million fragments using Samtools v1.7. The samples were down-sampled to 2.2 million fragments to meet the minimum whole genome sequencing depth requirement of ichorCNA (0.1X genome coverage). Multiple RBS of the same bam file with different seed was carried out to mimic shallow whole genome sequencing and to construct technical replicates so as to ensure that a particular seed did not confound the results. For ichorCNA analysis, we created different panels of normal for each feature and cutoff from the pool of controls to reduce noise and correct for systematic biases that may arise from library construction, sequencing platform, cfDNA specific artifacts, and feature selection. Lastly, we determined the importance of the feature with a given cutoff by calculating the tumor fraction of the 10 RBS with cutoff and no cutoff bam files using ichorCNA and their respective panel of 17ormal. This process will be referred to as the ctDNA in-silico bootstrap enrichment process (CISBEP). CISBEP was carried out for all 4 features individually and with combination of features with various cutoffs.

To determine which cutoff resulted in the highest enrichment of tumor fragments, we calculated the relative tumor fraction, which was calculated as the ratio of median tumor fraction from the 10 RBS of fragments within cutoff to the median tumor fraction of fragments with no cutoff.

### Effect of fragmentation feature selection on TP53 MAF

To further validate our findings, we measured the effect of these features on TP53 MAF. From the WGS data, we identified the sequenced fragments that covered the individual TP53 point mutation locus of HGSOC Patient 2-5 (Patient 1 had an insertion). Using a custom python script, we annotated each fragment as wildtype or mutant depending on whether it harbored the unique point mutation from the individual patients. We next annotated the fragments with fragment size and PBTNB-score. We did not investigate positioning of the fragments relative to the nucleosome center since some TP53 point mutations did not overlap any nucleosomes from CH01. We next selected fragments from HGSOC Patient 2-5 based on certain features, and calculated an aggregated TP53 MAF by dividing number of selected fragments that were mutant with total selected fragments. We only used single feature cutoffs that allowed the aggregated depth of at least 25. These features were: PBTNB-score < 0, PBTNB-score < -0.1, sizes 74-144bp, and sizes 74-144bp or 240-325bp. We next calculated a relative TP53 MAF by taking the ratio of the aggregated TP53 MAF with some feature selection to no feature selection.

## Data Availability

Anonymized data created for the study are or will be available in a persistent repository upon publication.

## Data and materials availability

Sequencing data from healthy volunteers and HGSOC patients for this study are deposited in the EGA database (accession numbers EGAS00001003258 and XXX for controls and cases respectively). Other data associated with this study are present in the paper.

## Acknowledgments

We would like to thank the Genomics and Bioinformatics core facilities at CRUK Cambridge Institute. We also acknowledge support from the Cancer Research UK Cambridge Institute, the NIHR Biomedical Research Centre, NIHR Cambridge Clinical Research Centre, and Experimental Cancer Medicine Centre. For their assistance with DNA extraction from plasma samples, we thank the Cancer Molecular Diagnostics Laboratory/Blood Processing Laboratory, which is supported by Cambridge NIHR Biomedical Research Centre, Cambridge Cancer Centre and the Mark Foundation of Cancer Research. Finally, we thank the patients and healthy volunteers for their contribution to this study. This study was supported by grants from CRUK Cambridge Institute (Core Grant, A29580) and by a funding from the ERC under the European Union’s Seventh Framework Programme (FP/2007-2013)/ERC Grant Agreement n. 337905.

## References

1. Wan JCM, Massie C, Garcia-Corbacho J, et al. Liquid biopsies come of age: towards implementation of circulating tumour DNA. Nat Rev Cancer. 2017;17(4):223–238. doi:10.1038/nrc.2017.7

2. Lennon AM, Buchanan AH, Kinde I, et al. Feasibility of blood testing combined with PET-CT to screen for cancer and guide intervention. Science (80-). April 2020:eabb9601. doi:10.1126/science.abb9601

3. Wan JCM, Heider K, Gale D, et al. ctDNA monitoring using patient-specific sequencing and integration of variant reads. Sci Transl Med. 2020;12(548):eaaz8084. doi:10.1126/scitranslmed.aaz8084

4. Bettegowda C, Sausen M, Leary RJ, et al. Detection of circulating tumor DNA in early-and late-stage human malignancies. Sci Transl Med. 2014;6(224):224ra24. doi:10.1126/scitranslmed.3007094

5. Mouliere F, Chandrananda D, Piskorz AM, et al. Enhanced detection of circulating tumor DNA by fragment size analysis. Sci Transl Med. 2018;10(466):eaat4921. doi:10.1126/scitranslmed.aat4921

6. Jiang P, Chan CWM, Chan KCA, et al. Lengthening and shortening of plasma DNA in hepatocellular carcinoma patients. Proc Natl Acad Sci. 2015;112(11):E1317–E1325. doi:10.1073/pnas.1500076112

7. Underhill HR, Kitzman JO, Hellwig S, et al. Fragment Length of Circulating Tumor DNA. PLoS Genet. 2016;12(7). doi:10.1371/journal.pgen.1006162

8. P J, K S, W P, et al. Plasma DNA End-Motif Profiling as a Fragmentomic Marker in Cancer, Pregnancy, and Transplantation. Cancer Discov. 2020;10(5):664–673. doi:10.1158/2159-8290.CD-19-0622

9. Jiang P, Sun K, Tong YK, et al. Preferred end coordinates and somatic variants as signatures of circulating tumor DNA associated with hepatocellular carcinoma. Proc Natl Acad Sci. 2018;115(46):E10925–E10933. doi:10.1073/PNAS.1814616115

10. Ulz P, Perakis S, Zhou Q, et al. Inference of transcription factor binding from cell-free DNA enables tumor subtype prediction and early detection. Nat Commun 2019 101. 2019;10(1):1–11. doi:10.1038/s41467-019-12714-4

11. Serpas L, Chan RWY, Jiang P, et al. Dnase1l3 deletion causes aberrations in length and end-motif frequencies in plasma DNA. Proc Natl Acad Sci. 2019;116(2):641–649. doi:10.1073/PNAS.1815031116

12. DSC H, M N, RWY C, et al. The Biology of Cell-free DNA Fragmentation and the Roles of DNASE1, DNASE1L3, and DFFB. Am J Hum Genet. 2020;106(2):202–214. doi:10.1016/J.AJHG.2020.01.008

13. Watanabe T, Takada S, Mizuta R. Cell-free DNA in blood circulation is generated by DNase1L3 and caspase-activated DNase. Biochem Biophys Res Commun. 2019;516(3):790–795. doi:10.1016/J.BBRC.2019.06.069

14. Moss J, Magenheim J, Neiman D, et al. Comprehensive human cell-type methylation atlas reveals origins of circulating cell-free DNA in health and disease. Nat Commun. 2018;9(1):5068. doi:10.1038/s41467-018-07466-6

15. Rostami A, Lambie M, Yu CW, Stambolic V, Waldron JN, Bratman S V. Senescence, Necrosis, and Apoptosis Govern Circulating Cell-free DNA Release Kinetics. Cell Rep. 2020;31(13):107830. doi:10.1016/j.celrep.2020.107830

16. Snyder MW, Kircher M, Hill AJ, Daza RM, Shendure J. Cell-free DNA Comprises an in Vivo Nucleosome Footprint that Informs Its Tissues-Of-Origin. Cell. 2016;164(1-2). doi:10.1016/j.cell.2015.11.050

17. Adalsteinsson VA, Ha G, Freeman SS, et al. Scalable whole-exome sequencing of cell-free DNA reveals high concordance with metastatic tumors. Nat Commun. 2017;8(1). doi:10.1038/s41467-017-00965-y

18. Sun K, Jiang P, Wong AIC, et al. Size-tagged preferred ends in maternal plasma DNA shed light on the production mechanism and show utility in noninvasive prenatal testing. Proc Natl Acad Sci. 2018;115(22):E5106–E5114. doi:10.1073/PNAS.1804134115

19. Kaplan N, Moore IK, Fondufe-Mittendorf Y, et al. The DNA-encoded nucleosome organization of a eukaryotic genome. Nat 2008 4587236. 2008;458(7236):362–366. doi:10.1038/nature07667

20. Chandrananda D, Thorne NP, Bahlo M. High-resolution characterization of sequence signatures due to non-random cleavage of cell-free DNA. BMC Med Genomics. 2015;8(1):29. doi:10.1186/s12920-015-0107-z

21. Cristiano S, Leal A, Phallen J, et al. Genome-wide cell-free DNA fragmentation in patients with cancer. Nature. May 2019:1. doi:10.1038/s41586-019-1272-6

22. Forshew T, Murtaza M, Parkinson C, et al. Noninvasive Identification and Monitoring of Cancer Mutations by Targeted Deep Sequencing of Plasma DNA. Sci Transl Med. 2012;4(136):136ra68–136ra68. doi:10.1126/scitranslmed.3003726

