## supplementary_figures for "Refined characterization of circulating tumor DNA through biological feature integration"

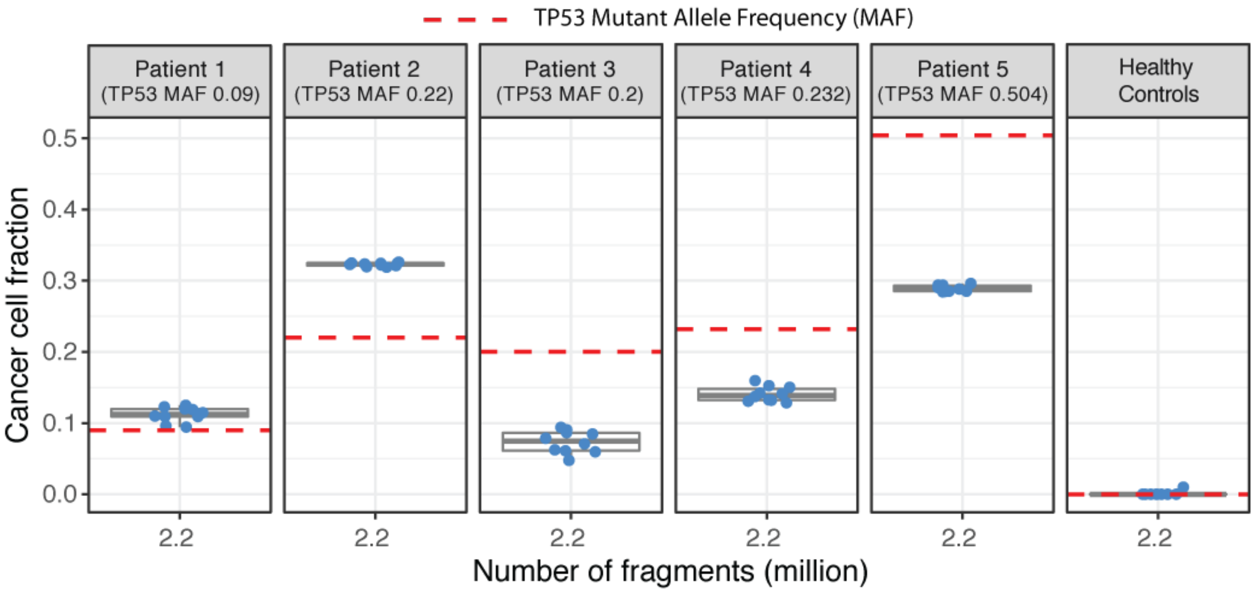


**Supplementary Figure 1.** The cancer cell fraction (CCF) as estimated by copy number quantification analysis of 10 replicates from each patient and control panel. The red dashed horizontal line marks the TP53 mutant allele fraction measured using targeted amplicon sequencing.

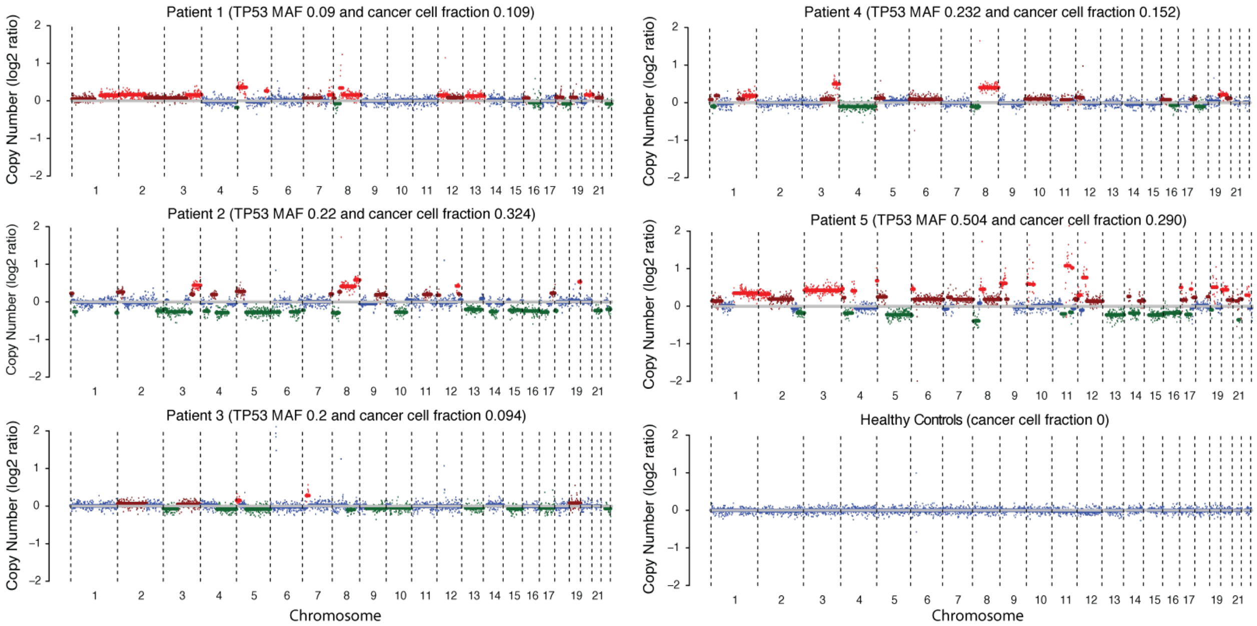


**Supplementary Figure 2.** Copy number profiles of the 5 HGSOC patients and panel of healthy controls before any feature selection using shallow whole-genome sequencing at 0.1x coverage (2.2 million fragments).

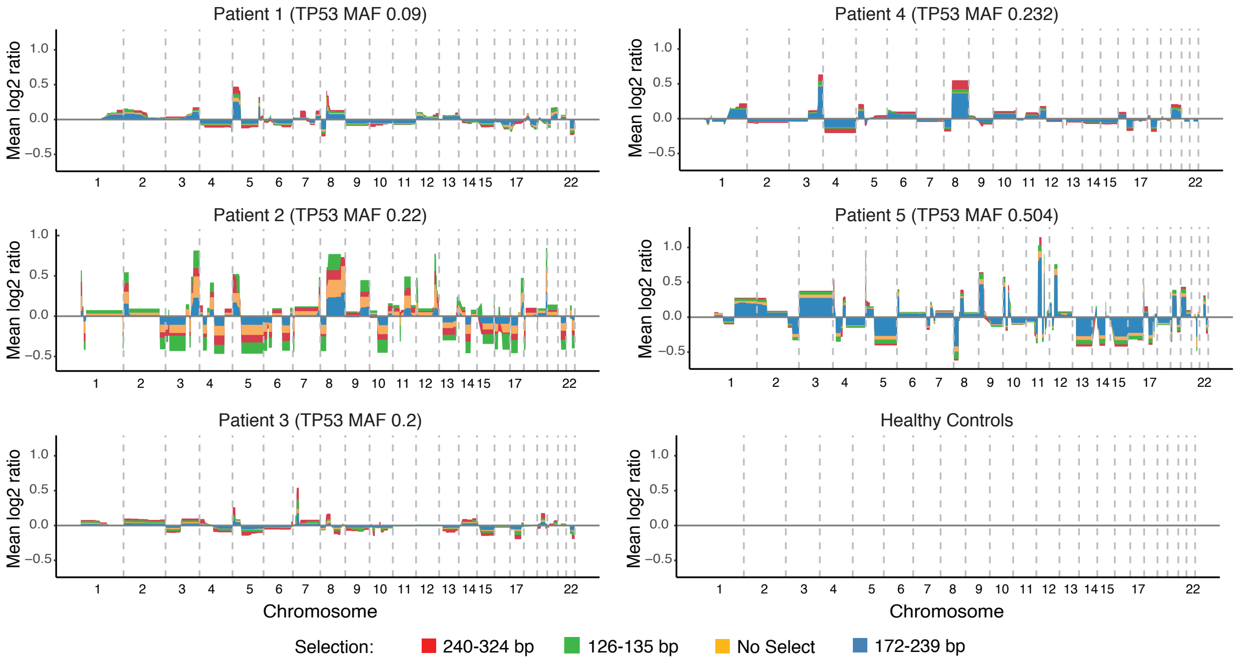


**Supplementary Figure 3.** The average relative copy number values from the 10 replicates of each patient. The different colors indicate results from using fragments with no size selection, and those with fragment length 240-324bp and 172-239bp.

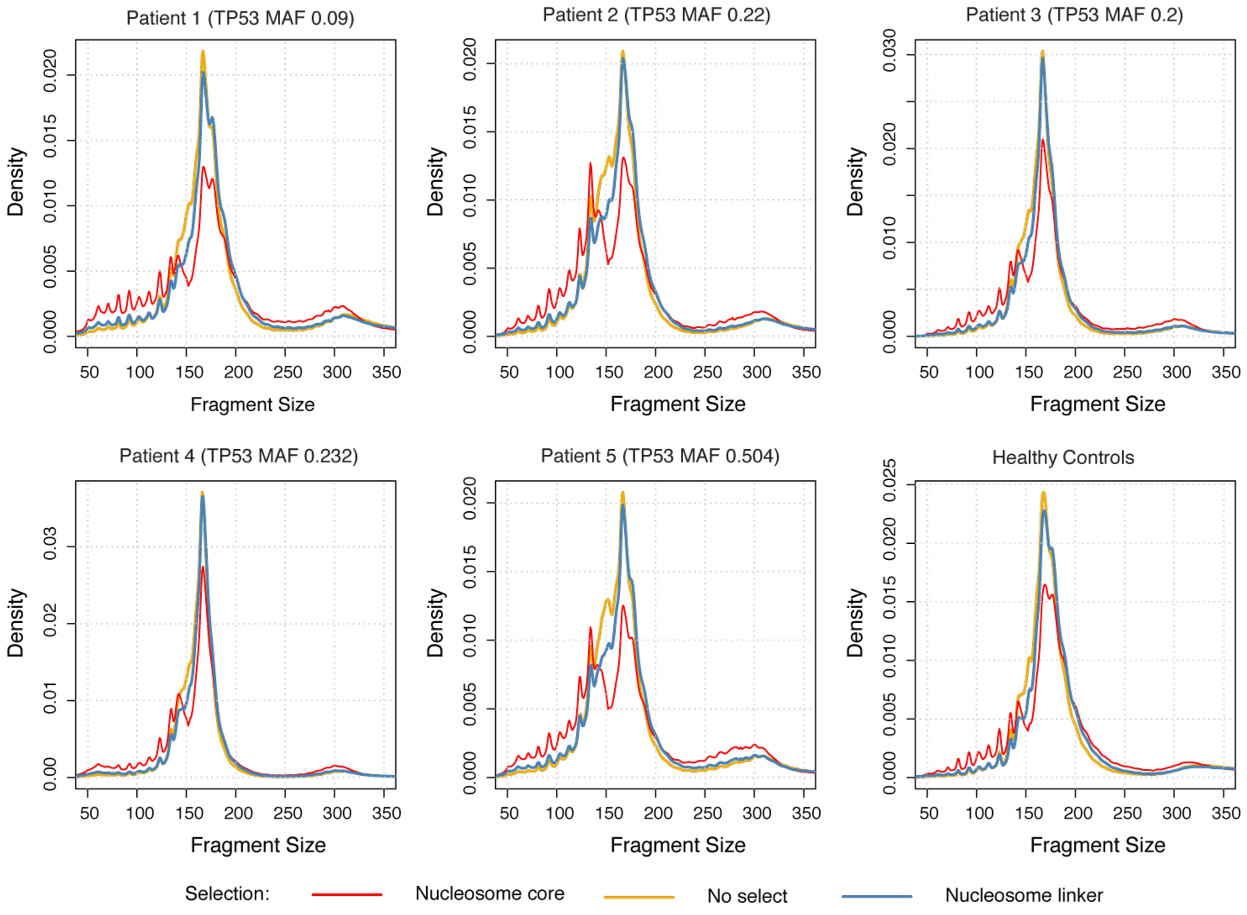


**Supplementary Figure 4.** Fragment size distribution of fragments with no size selection, fragments that start and end within nucleosome core region, and fragments that start and end within nucleosome linker region.

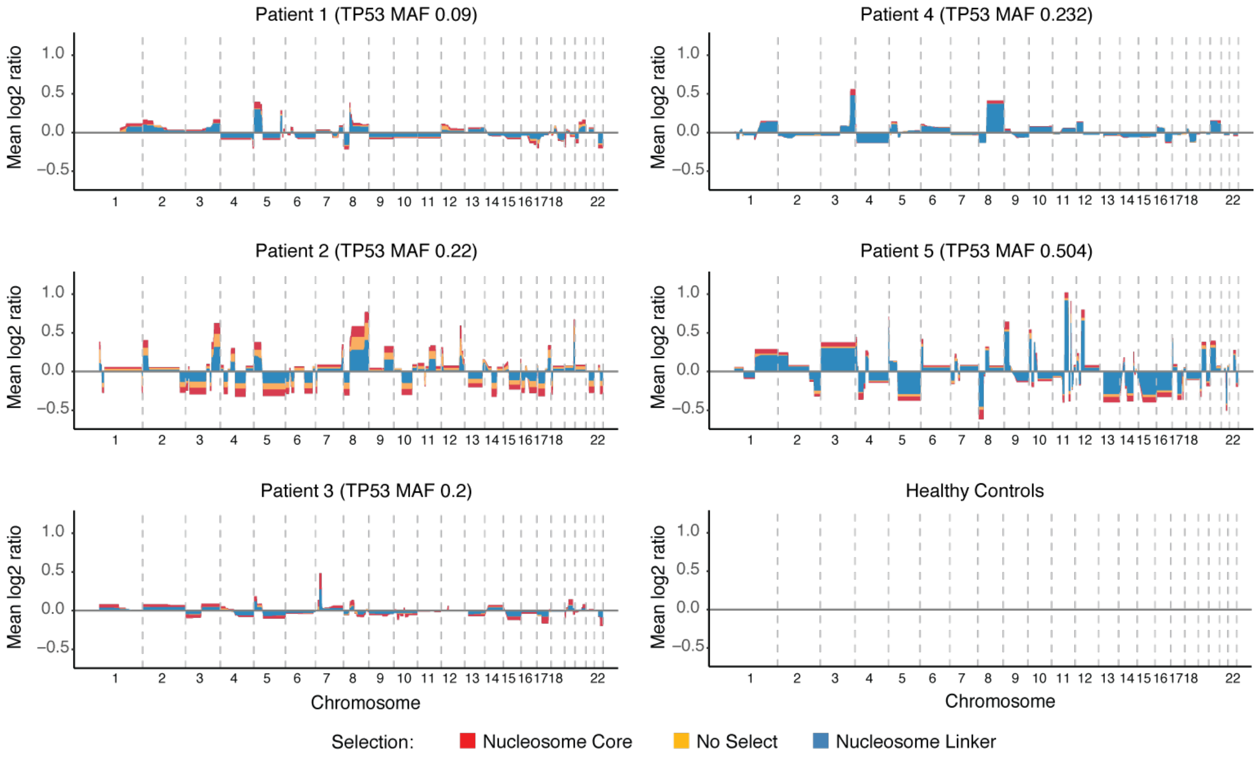


**Supplementary Figure 5.** The average relative copy number values from the 10 replicates of each patient. The different colors indicate results from using fragments with no size selection, fragments that start and end within nucleosome core region, and fragments that start and end within nucleosome linker region.

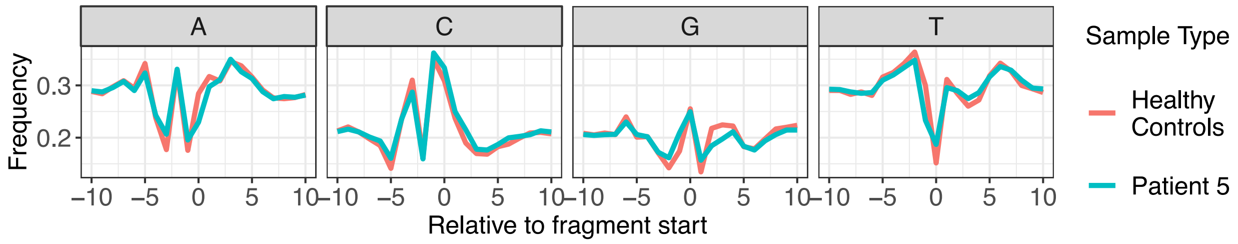


**Supplementary Figure 6.** Per base mean mono-nucleotide frequency of 10bp downstream and upstream of fragment start sites. Calculated on a set of 3 million fragments randomly selected 10 times from the panel of healthy controls and HGSOC Patient 5.

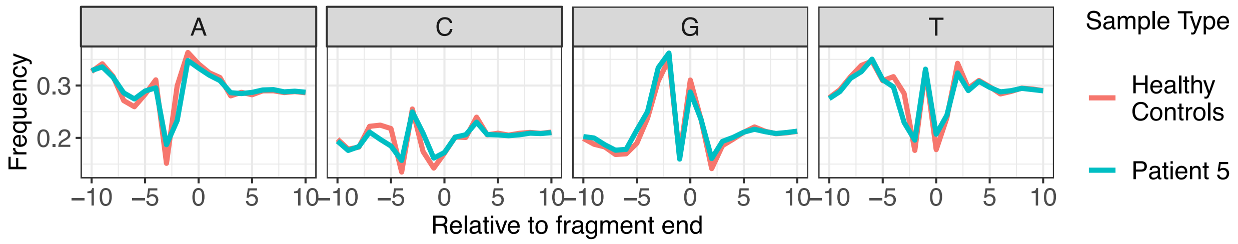


**Supplementary Figure 7.** Per base mean mono-nucleotide frequency of 10bp downstream and upstream of fragment end sites. Calculated on a set of 3 million fragments randomly selected 10 times from the panel of healthy controls and HGSOC Patient 5.

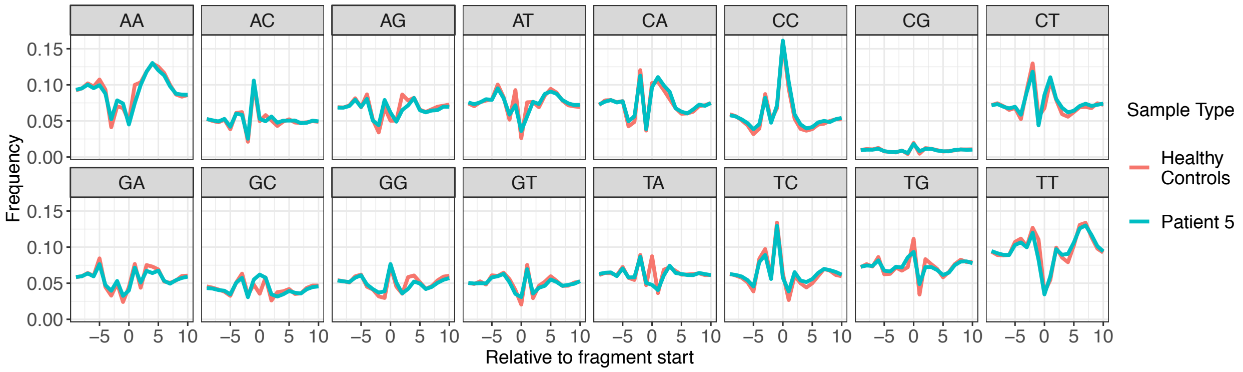


**Supplementary Figure 8.** Per base mean di-nucleotide frequency of 10bp downstream and upstream of fragment start sites. Calculated on a set of 3 million fragments randomly selected 10 times from the panel of healthy controls and HGSOC Patient 5.

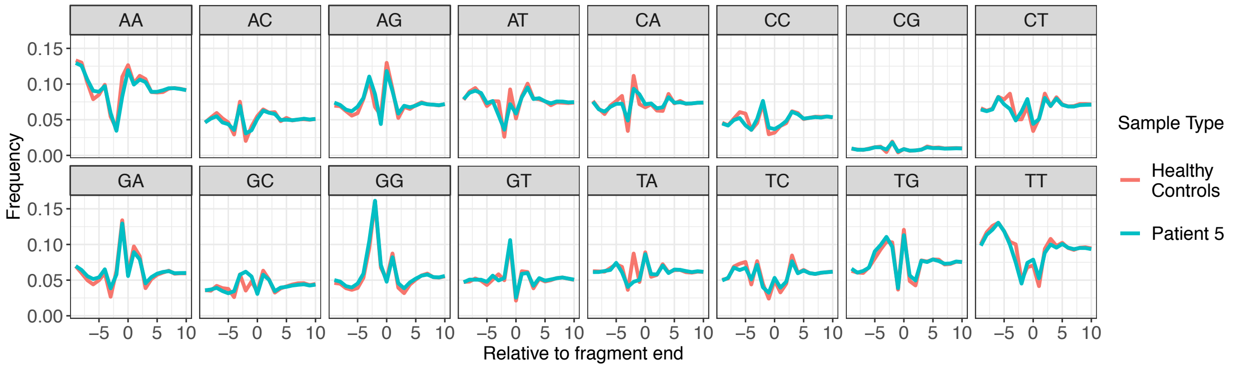


**Supplementary Figure 9.** Per base mean di-nucleotide frequency of 10bp downstream and upstream of fragment end sites. Calculated on a set of 3 million fragments randomly selected 10 times from the panel of healthy controls and HGSOC Patient 5.

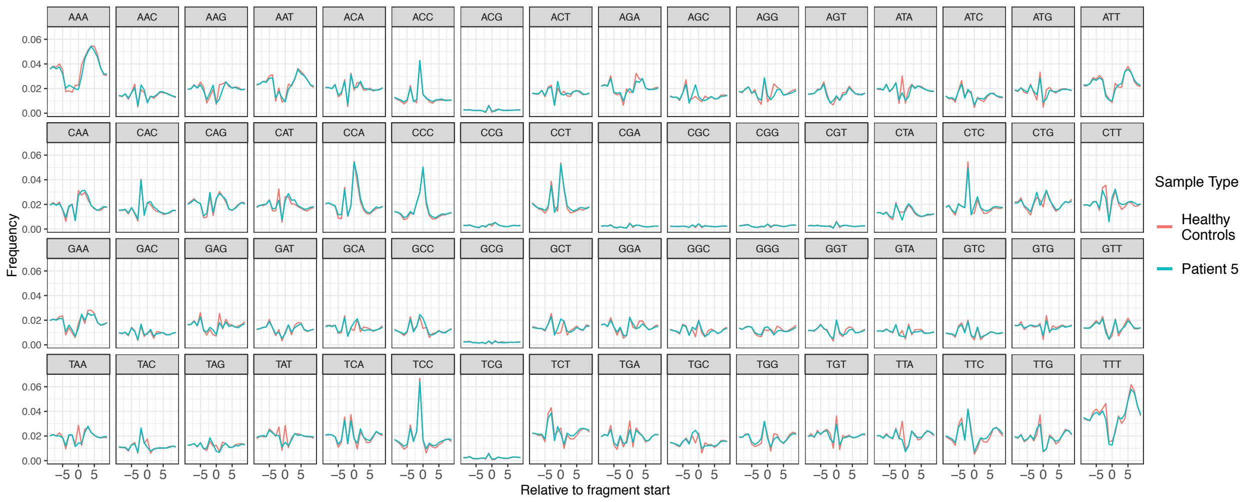


**Supplementary Figure 10.** Per base mean tri-nucleotide frequency of 10bp downstream and upstream of fragment start sites. Calculated on a set of 3 million fragments randomly selected 10 times from the panel of healthy controls and HGSOC Patient 5.

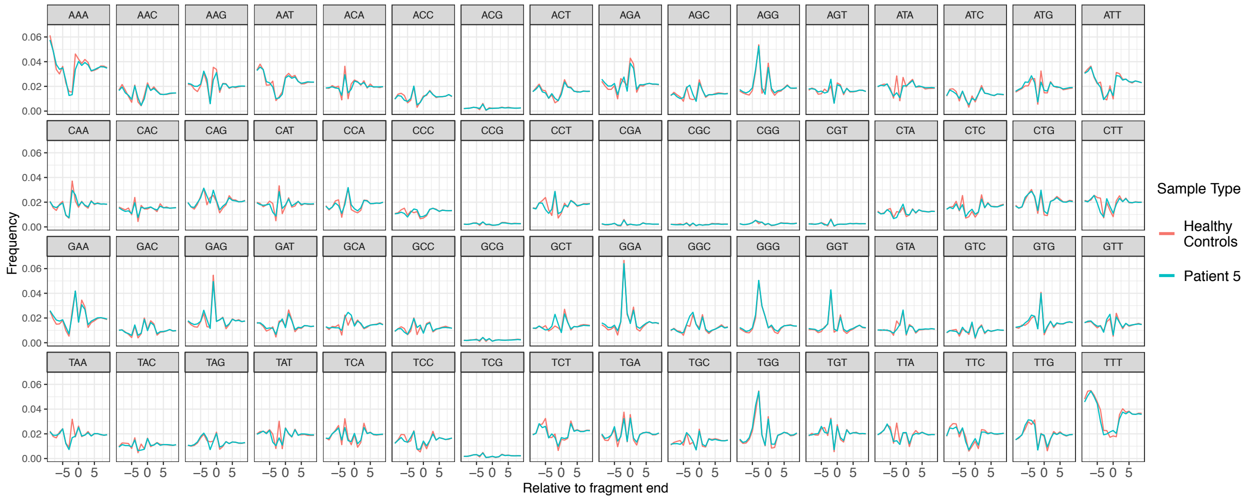


**Supplementary Figure 11.** Per base mean tri-nucleotide frequency of 10bp downstream and upstream of fragment end sites. Calculated on a set of 3 million fragments randomly selected 10 times from the panel of healthy controls and HGSOC Patient 5.

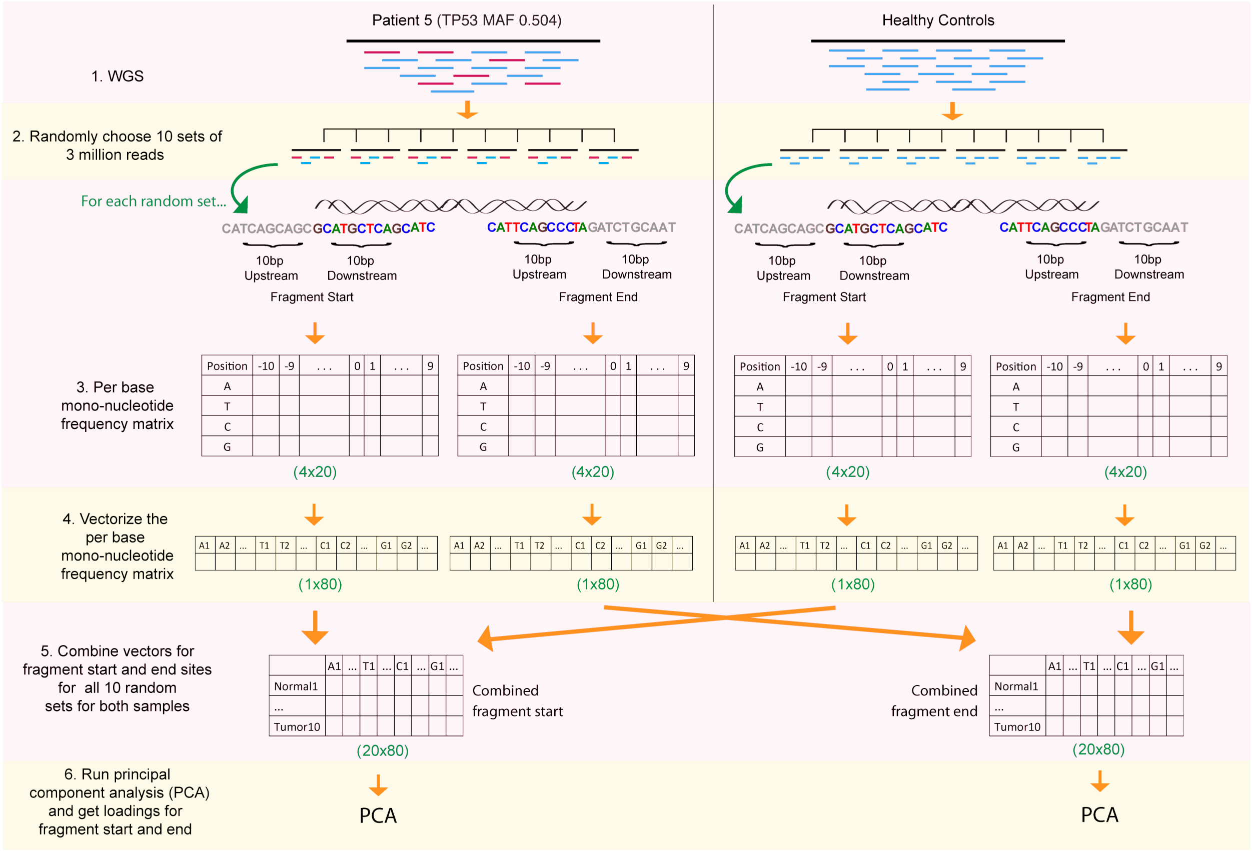


**Supplementary Figure 12.** Schematic representation of the first phase to calculate the mono-nucleotide frequency score. Similar steps were taken to calculate the di- and tri-nucleotide frequency scores.

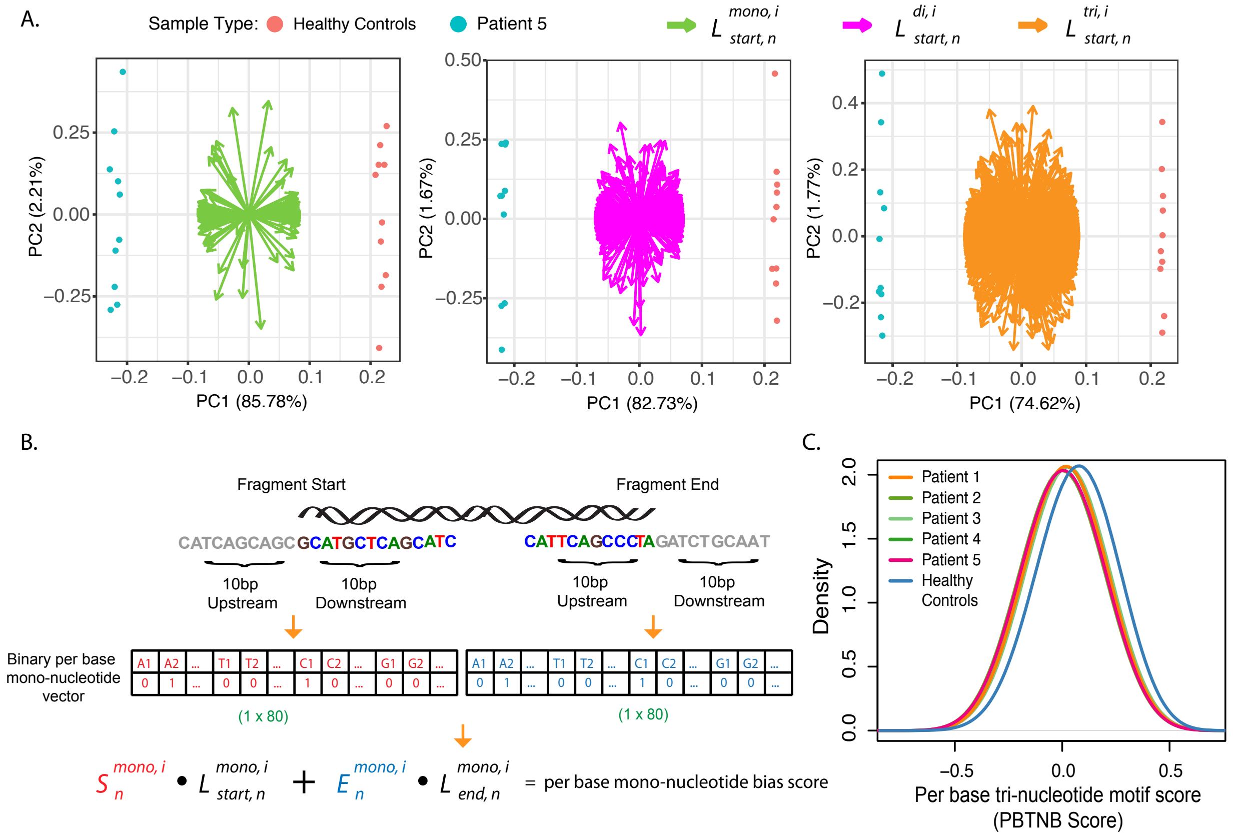


**Supplementary Figure 13.** Steps to calculate nucleotide motif scores. (A) 10 technical replicates were created for the panel of healthy controls and Patient 5 by randomly selecting 3 million fragments. For each replicate, 6 position weight matrices (PWM) were constructed describing the per base mono-, di-, and tri- nucleotide frequencies of the genomic sequences flanking 10bp downstream and upstream of both fragment start and end sites. Principal component analysis was then carried out on the collapsed and concatenated mono-, di-, and tri- nucleotide PWM from all 10 technical replicates of healthy controls and Patient 5 (See Supplementary Figure 4 for more details). All replicates from the panel of healthy controls and Patient 5 clustered separately in all 3 matrices that summarized the mono-, di-, and tri- nucleotide frequencies of fragment start sites. Data for fragment end sites are not shown, however similar clustering of both sample types was observed. The loading vectors of fragment start sites for mono-, di-, and tri- nucleotide frequencies are also shown in each plot. (B) Schematic representation of how to calculate the mono-nucleotide motif score for an individual fragment using the principal component loading vectors of fragment start and end site mono-nucleotide frequencies. For a given fragment, two binary per base mono-nucleotide fragments are created that indicate whether a given nucleotide is present at a given position of fragment start and end site. Then the mono-nucleotide motif score is calculated by the sum of the dot product of binary mono-nucleotide vector of fragment start and end site with the principal component loading vector of fragment start and end site mono-nucleotide frequencies. Similar steps were taken to calculate the di- and tri- nucleotide motif scores. (C) The distribution of per base tri-nucleotide bias score for all fragments of 5 HGSOC patients and panel of healthy controls.

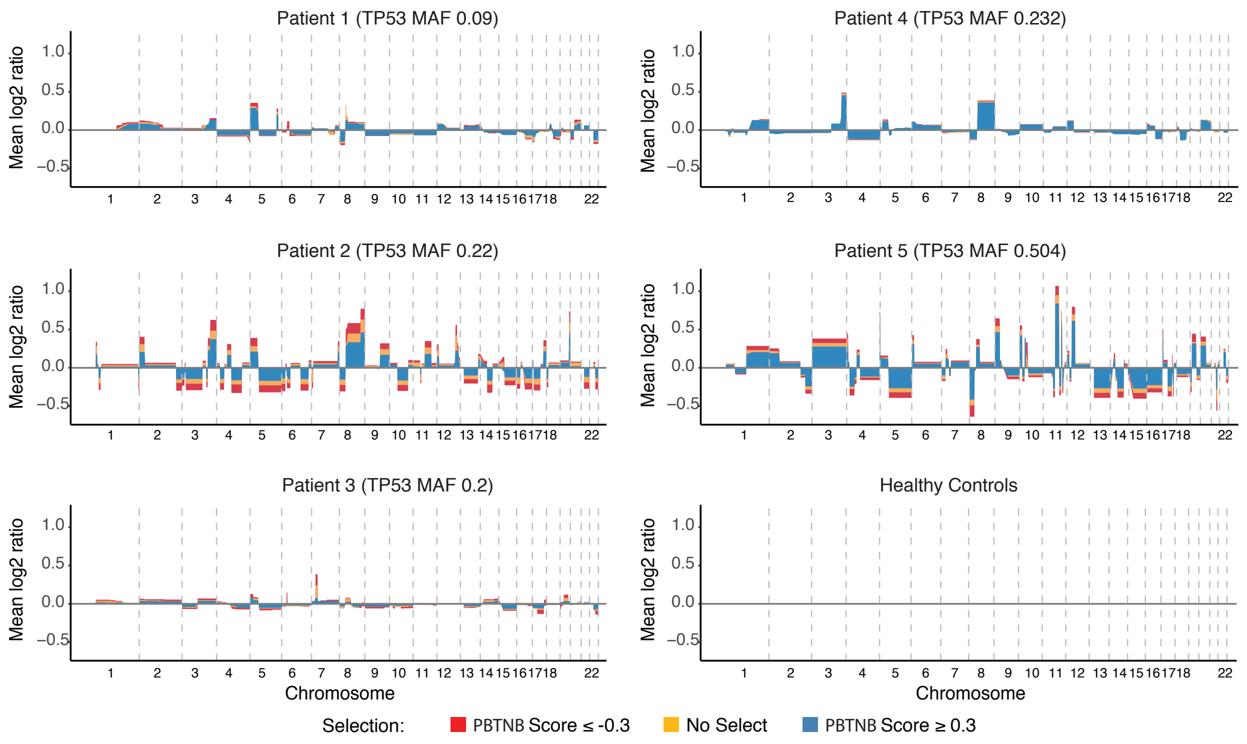


**Supplementary Figure 14.** The average relative copy number values from the 10 replicates of each patient. The different colors indicate results from fragments with no selection, fragments with per base tri-nucleotide bias score (PBTNB score) less than or equal to -0.3 and greater than or equal to 0.3.

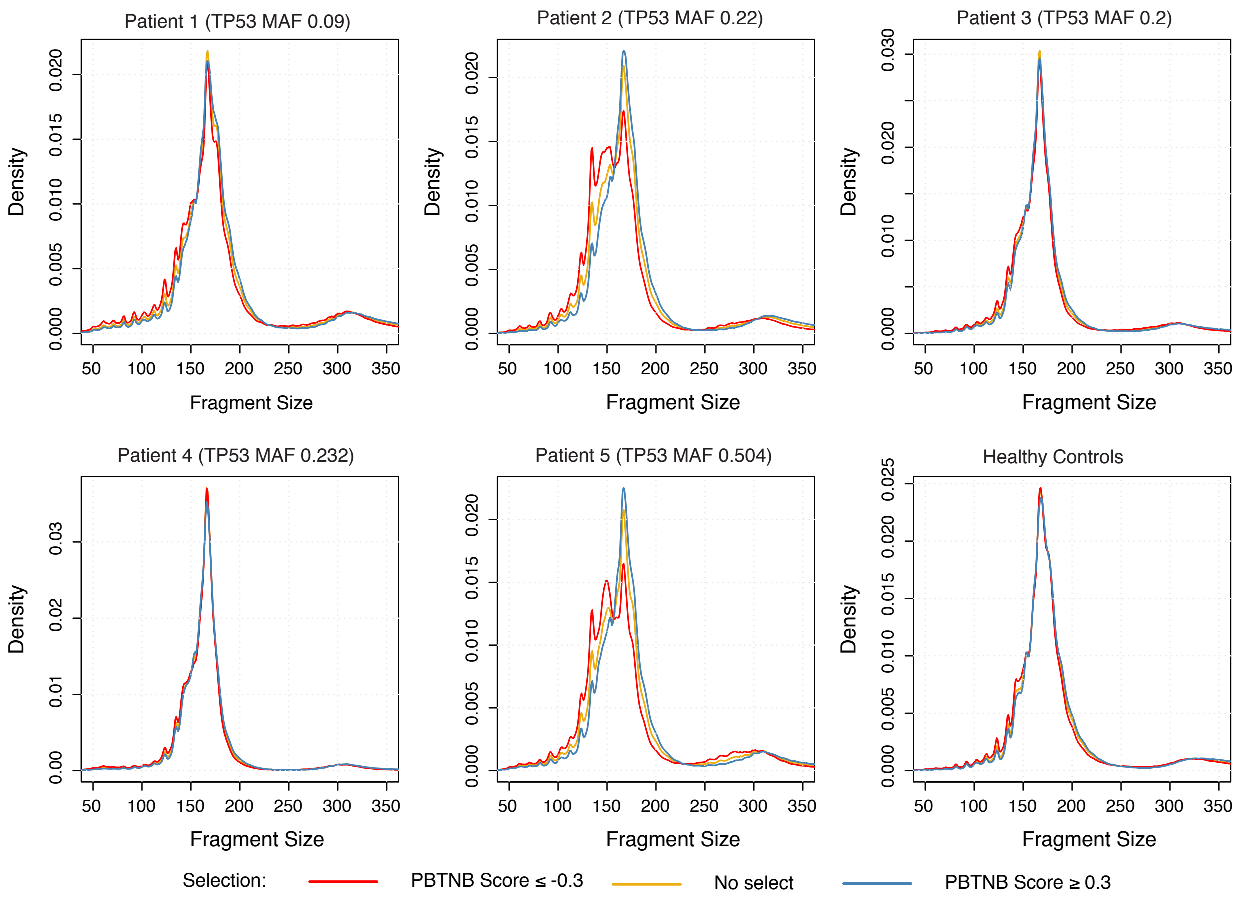


**Supplementary Figure 15.** Fragment size distribution of fragments with no size selection, fragments with tri-nucleotide motif score less than or equal to -0.3 and greater than or equal to 0.3.
